# Phylogeography of hepatitis B: The role of Portugal in the early dissemination of HBV worldwide

**DOI:** 10.1101/2022.01.05.22268725

**Authors:** Rute Marcelino, Ifeanyi Jude Ezeonwumelu, André Janeiro, Paula Mimoso, Sónia Matos, Veronica Briz, Victor Pimentel, Marta Pingarilho, Rui Tato Marinho, José Maria Marcelino, Nuno Taveira, Ana Abecasis

**Affiliations:** Instituto Universitário Egas Moniz, Monte de Caparica, Portugal; Research Institute for Medicines (iMed.ULisboa), Faculty of Pharmacy, Universidade de Lisboa, Lisbon, Portugal; Global Health and Tropical Medicine (GHTM), Instituto de Higiene e Medicina Tropical/Universidade Nova de Lisboa (IHMT/UNL), Lisbon, Portugal; AIDS Research Institute-IrsiCaixa and Health Research Institute Germans Trias i Pujol (IGTP), Hospital Germans Trias i Pujol, Universitat Autònoma de Barcelona, Badalona, Spain; GenoMed - Diagnósticos de Medicina Molecular, Instituto de Medicina Molecular, Faculdade de Medicina de Lisboa, Portugal; Laboratory of Viral Hepatitis, National Center for Microbiology, Institute of Health Carlos III, Madrid, Spain; Department of Gastroenterology and Hepatology, Santa Maria Hospital, Medical School of Universidade de Lisboa, Lisbon, Portugal

## Abstract

In 2015, 257 million people lived with chronic hepatitis B infection. In Portugal 1.4% of the population is chronically infected with hepatitis B virus. Limited information is available regarding the genetic diversity, origin of HBV in Portugal and its role in the dissemination of HBV worldwide. In this study, we aimed to investigate the epidemic history and transmission dynamics of HBV genotypes that are endemic in Portugal. HBV *pol* gene was sequenced from 130 patients followed in Lisbon. HBV genotype A (HBV/A) was the most prevalent genotype (n=54, 41.5%), followed by D [HBV/D; (n=44, 33.8%)], and E [HBV/E; (n=32, 24.6%)]. Spatio-temporal evolutionary dynamics were reconstructed using a Bayesian Markov Chain Monte Carlo method, as implemented in BEAST v1.10.4, with a GTR nucleotide substitution model, an uncorrelated lognormal relaxed molecular clock model under a Bayesian skyline plot and a continuous diffusion model. Our results indicate that HBV/D4 was the first subgenotype to be introduced in Portugal by the end of the XIX century, around 1857 (HPD 95% 1699 - 1931) followed by subgenotypes D3 and A2 a few decades later. Genotype E and subgenotype A1 were introduced in Portugal later, almost simultaneously. Our results also indicate a very important role of Portugal in the exportation of HBV subgenotypes D4 and A2 to Brazil and Cape Verde, respectively, in the beginning of the XX century. This work clarifies the epidemiological history of HBV in Portugal and show that Portugal had an important role in the global spread of this virus.

## Introduction

Hepatitis B virus (HBV) infection is a severe life-threatening disease with an estimated prevalence, in 2015, of 257 million infections worldwide and a reported death toll of 887,000 deaths due to complications related to HBV chronic infection, such as liver cirrhosis and hepatocellular carcinoma (1). Despite the existence of an effective vaccine since 1982 and potent antiviral drugs HBV infection remains a serious public health concern (1).

HBV belongs to the *Hepadnaviridae* family and like all the viruses from this family it causes a hepatotropic infection that drives a lifelong dynamic disease, where the immunological response of the host against the virus induces an ongoing liver inflammation process. This inflammation increases the risk of fibrosis, cirrhosis, end-stage liver disease and hepatocellular carcinoma (2). The clinical outcomes may vary due to different host and viral factors, such as gender, age, ethnicity, quantitative HBs antigen (HBsAg) levels, viral load, specific mutations of the virus and HBV genotype (3-5).

There are nine viral genotypes (A to I), having a pairwise genomic divergence of at least 7.5%. Genotypes comprise several subgenotypes having a pairwise divergence of 4% to 7.5%. There are three A subgenotypes A1, A2, A4, and quasisubgenotype A3, six B subgenotypes B1, B2, B4–B6, and quasisubgenotype B3, six C subgenotypes C1 – C16, six D subgenotypes D1–D6, four F subgenotypes F1-F4 and two I subgenotypes I1 and I2 (6). A potential 10^th^ genotype, genotype J, was found to be a recombinant of genotype C and gibbon HBV in the S region and that may represent an independent cross-species transmission (7).

The high HBV genetic diversity occurs due to the lack of proofreading activity of the viral reverse transcriptase (RT) enzyme used in its replication process. This makes the virus evolve extremely fast, with an estimated evolutionary rate of about 10^−4^-10^−6^ nucleotide substitutions/site/year (s/s/y). This represents an evolutionary rate around 100 times higher than any other DNA virus (5).

In Europe, vaccination campaigns seem to have contributed to a decline in HBV prevalence (8). Portugal is a low endemicity country with an HBsAg prevalence in the general population that decreased along the years until achieving 0.02–1.45% in the decades of 1990-2014 (9-11). Nonetheless, 2.9% of hospital admissions for liver cirrhosis in Portugal, between 2003 and 2012, were related to hepatitis B (12).

Limited information exists regarding HBV diversity in Portugal. Between 2004 and 2014 genotype D was the most prevalent genotype in chronically-infected Portuguese patients (9). Genotype E was referred as more common in Central-Southern (10-62%) than in Northern Portugal (1-4%) (9, 13). Genotypes C and F have been reported in the North of the country on rare occasions (14). In addition to this, previously unpublished data of a small patient population of Hospital Santa Maria in Lisbon indicated the circulation of minority genotypes such as B, C and F (15). Historical migration patterns to and from Portugal may have shaped the epidemiological profile of HBV infection in the country and, therefore, it is important to investigate the epidemiological history, transmission dynamics and origin of HBV strains circulating in Portugal. The aim of this study was to characterize the epidemic history of HBV genotypes in Portugal as well as understanding Portugal’s role in the spread of HBV around the world, through the reconstruction of the spatiotemporal evolutionary dynamics of the virus using a Bayesian framework.

## Materials and Methods

### Study participants and ethics statement

In this retrospective study we used plasma samples collected for diagnostic purposes from 2005 to 2012 from chronic HBV Portuguese patients assisted in Department of Gastroenterology and Hepatology of Santa Maria Hospital, Medical School of Lisbon, Portugal. Samples were selected for sequencing based on sample volume and the availability of information on age and gender. This study was approved by the Institutional Review Board of Santa Maria Hospital (ref. 600/15 of January 7^th^, 2016) and followed all ethical principles of the Declaration of Helsinki.

### HBV genotyping

Viral nucleic acid was extracted from 200 µl of plasma using the QIAamp DNA Blood Mini Kit (Qiagen, USA). A 749 base pair (bp) fragment of the *pol* gene region was amplified by nested PCR with the BIOTAQ DNA polymerase (Bioline, United Kingdom) using the following primer pairs (positions based on the complete reference sequence NC_003977): 5’-ATCCTCACAATACCGCAGA-3’ (position: 229-247) and 5’-AGGAGTTCCGCAGTATGG-3’ (position: 1285-1268) (First round); 5’-AGACTCGTGGTGGACTTCTCT-3’ (position: 252-272) and 5’-GCGTCAGCAAACACTT-3’ (position: 1195-1180) (Second round). The thermocycling conditions for both PCR rounds were: 1 cycle at 94ºC for 4 min, and 40 cycles at 94ºC for 45 sec; 55ºC for 30 sec; 72ºC, 1 min with increments of 5 sec/cycle. A final elongation step of 15 min at 72ºC was included at the end of amplification run. Amplified DNA products were sequenced on the automated sequencer ABI-3130XL (Applied Biosystems, USA) using the second PCR round primer pair and BigDye terminator v. 3.1 cycle sequencing kit (Applied Biosystems, USA). The sequences were analyzed using Chromas Pro.1.7.6 software (Technelysium, Pty, Australia). Multiple sequence alignment was performed using the ClustalW algorithm, as implemented in the software ClustalX 2.1 (16). Genomic sequences were genotyped using the Geno2Pheno HBV online tool (https://hbv.geno2pheno.org/). The reliability of the Geno2Pheno results was confirmed by analyzing the consistency of its results with the results of phylogenetic analysis.

### Genetic Distances

Evolutionary distances between and within groups were estimated using the Kimura 2-parameter evolutionary model, by calculating the average evolutionary distance between all pairs of groups and between all pairs of sequences within groups, respectively. Standard error estimates were obtained using a bootstrap procedure (1000 replicates). All analyses were conducted in MEGA7 (17). Rate variation among sites was modelled with a gamma distribution (shape parameter = 1).

### Control sequences selection for spatiotemporal analysis

Control sequences for the spatiotemporal analysis were selected using the NCBI Nucleotide Blast (https://blast.ncbi.nlm.nih.gov/) algorithm. There were 9489 HBV complete genome and 6561 HBV pol gene sequences available. The alignment with the 130 Portuguese HBV sequences was submitted to NCBI nucleotide blast, selecting a maximum target of 50 sequences in the algorithm parameters. Thus, for each of the Portuguese sequences, we retrieved the 50 most similar HBV sequences available in Genbank, making a total of 6500 HBV sequences for the control group. Multiple sequence alignment was performed on this group of 6500 sequences using ClustalW, as implemented in the ClustalX 2.1 software (16). Duplicate sequences were removed from this dataset with Python v3.7.0 (https://www.python.org/downloads/release/python-370/), reducing the data file to 687 HBV control sequences. From this group, 286 sequences without collection date and country of origin were excluded, creating a data set with 401 control sequences. We tried to keep the temporal and spatial information as wide as possible in the control sequence dataset, diversifying as much as possible both the year of harvest and the country of origin of the control sequences. A manual search in the NCBI was carried out, selecting the sequences by genotype and year of harvest and/or country in an attempt to increase the representativeness of the 60s, 70s and 80s for most genotypes. We also had to increase the representativeness of the origin countries of A1 and D4 subgenotypes, to ensure that Brazil was not falsely over-represented in alignment since the result of the initial BLAST nucleotide resulted in near 35% of sequences from that country. With this approach, the final dataset consisted of 453 HBV control sequences spanning from 1963 to 2018 (supplementary table 1), and 130 Portuguese HBV sequences.

### Spatiotemporal evolutionary dynamics

To minimize the effect of convergent evolution, nine codon positions associated with treatment selective pressure were removed from the alignment. Time-scaled phylogeny, evolutionary rate and phylogeography were estimated using a Bayesian Markov Chain Monte Carlo (MCMC) method implemented in the BEAST v1.10.4 (18), with the GTR+F+R4 nucleotide substitution model that was selected using Model Finder module (19) within iqtree 1.6.11 software and the Akaike information criterion (20).

Six different evolutionary models were tested: strict vs relaxed molecular clock, each one combined with either parametric (expansion Growth and exponential Growth) and non-parametric (Bayesian Skygrid and Bayesian Skyline) demographic priors and with a continuous Cauchy RRW phylogeography model. In each case, two independent MCMC chains were run for 300 million generations (sampled every 30.000 steps) to ensure convergence and Effective Sample Size (ESS) values *>* 100. Our data only converged and showed acceptable ESS values when using an uncorrelated lognormal relaxed molecular clock model under a Bayesian skyline plot (a non-parametric piecewise-constant model) as coalescent priors.

Uncertainty of parameter estimates was assessed after excluding the initial 10% of the run by calculating the 95% Highest Probability Density (HPD) values using TRACER v1.7.1 program (21). Maximum clade credibility (MCC) tree was summarized from the posterior distribution of trees with TreeAnnotator included in BEAST package and visualized and annotated with FigTree v1.4.4 (http://tree.bio.ed.ac.uk/software/figtree/) to show node support values (posterior state probability) and median time of MRCAs with High Posterior Density 95% for each common ancestral node. Bifurcating nodes with posterior probability greater than 0.90 were considered statistically well supported.

## Results

### HBV genotype A was the most prevalent in Portugal

Most (n=97, 74.6%) of the patients included in the study were male with a median age of 43.6 years. Median age of females (n= 33, 25.4%) was 41.2 years. According to Geno2Pheno genotype classificatin, HBV genotype A (HBV/A) was the most prevalent genotype (n=54, 41.5%), followed by D [HBV/D; (n=44, 33.8%)], and E [HBV/E; (n=32, 24.6%)]. Subgenotypes A1 (HBV/A1) and D4 (HBV/D4) were almost equally prevalent with 23.1% (n=30) and 22.3% (n=29), respectively, followed by A2 (HBV/A2) with 16.2% (n=21) and D3 (HBV/D3) with 11.5% (n=15). Three sequences (n=2.3%) were classified as genotype A without further subgenotype classification.

Genetic distances between HBV genotypes and subgenotypes present in Portugal were >7.5% for genotypes and between 4% - 7.5% for subgenotypes, consistent with rules of classification and further validating the results obtained with the Geno2Pheno algorithm (Table 1).

**Table 1.**
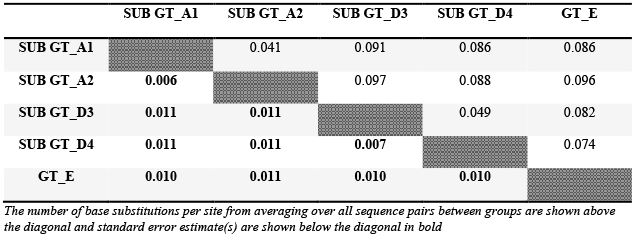
Divergence between HBV genotypes/subgenotypes present in Portugal.

Genetic divergence within genotypes/subgenotypes was higher for HBV/A1 and HBV/D3 and lower for HBV/E (Table 2).

**Table 2.**
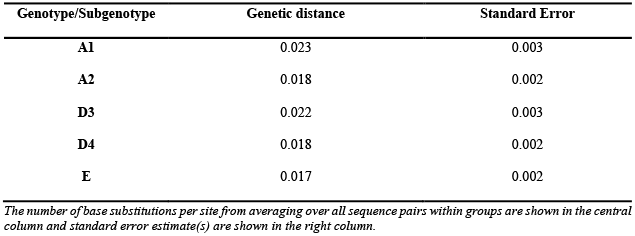
Divergence within HBV genotypes/subgenotypes isolates present in Portugal.

### HBV was first introduced in Portugal around 1857 via genotype D

The estimated mean substitution rate obtained in TRACER’s analysis was 9.45×10^−5^ s/s/y (95% HPD: 4.22×10^−5^ - 1.43×10^−4^). Data from spatiotemporal reconstruction analyses using this substitution rate suggest that HBV might have originated in France (posterior state probability (*PSP*) = 1) around the year 886 (95% HPD: 414 B.C - 1514) (Table 3). Among the studied genotypes, HBV/D was probably the first genotype to emerge in the beginning of XVI century, immediately followed by HBV/A. HBV/E was the last to emerge, maybe three centuries later than A (Table 3).

**Table 3.**
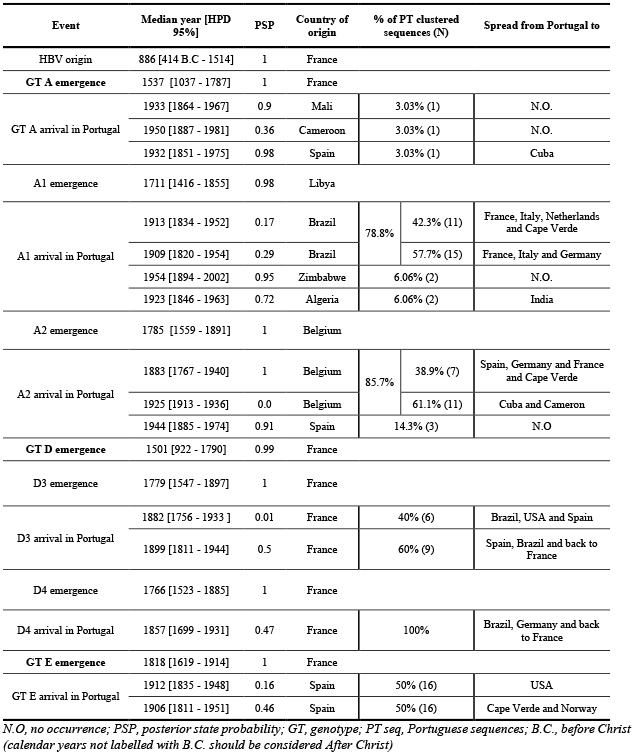
Estimated time and place of HBV origin, origin of the Portuguese strains, time of introduction in Portugal and role of Portugal in the global HBV spread.

In Portugal, HBV/D4 was the first to emerge possibly due to a single introduction estimated to have occurred around 1857 (95% HPD: 1699 – 1931). HBV/D3 and HBV/A2 were introduced around 1882 (95% HPD: 1756 – 1933) and 1883 (95% HPD: 1767 – 1940), respectively. HBV/D3 was introduced again in Portugal at the end of XIX century and HBV/A2 was again introduced in Portugal twice during the first half of XX century. HBV/A1 was introduced multiple times in Portugal, the first one having occurred near 1909 (95% HPD: 1820 – 1954) and the last three until 1954 (95% HPD: 1894 – 2002). Finally, the first and second introductions of HBV/E in Portugal, occurred six years apart in 1906 (95% HPD: 1811 – 1951) and 1912 (95% HPD: 1835 – 1948) (Table 3).

### Most HBV genotypes were imported to Portugal from other European countries

HBV seems to have evolved in its original region, France, during several years, diverging into one branch that gave rise to the present genotype A, and another one that was at the origin of the remaining genotypes studied (Figure 1). In Portugal, HBV/D3 and HBV/D4 were both introduced via France. The emergence of HBV/A2 in Portugal occurred via Belgium and Spain, while HBV/E was exclusively introduced in Portugal via Spain. The unique subgenotype that entered in Portugal via non-European countries was HBV/A1 (Figure 1; Table 3).

**Figure 1.**
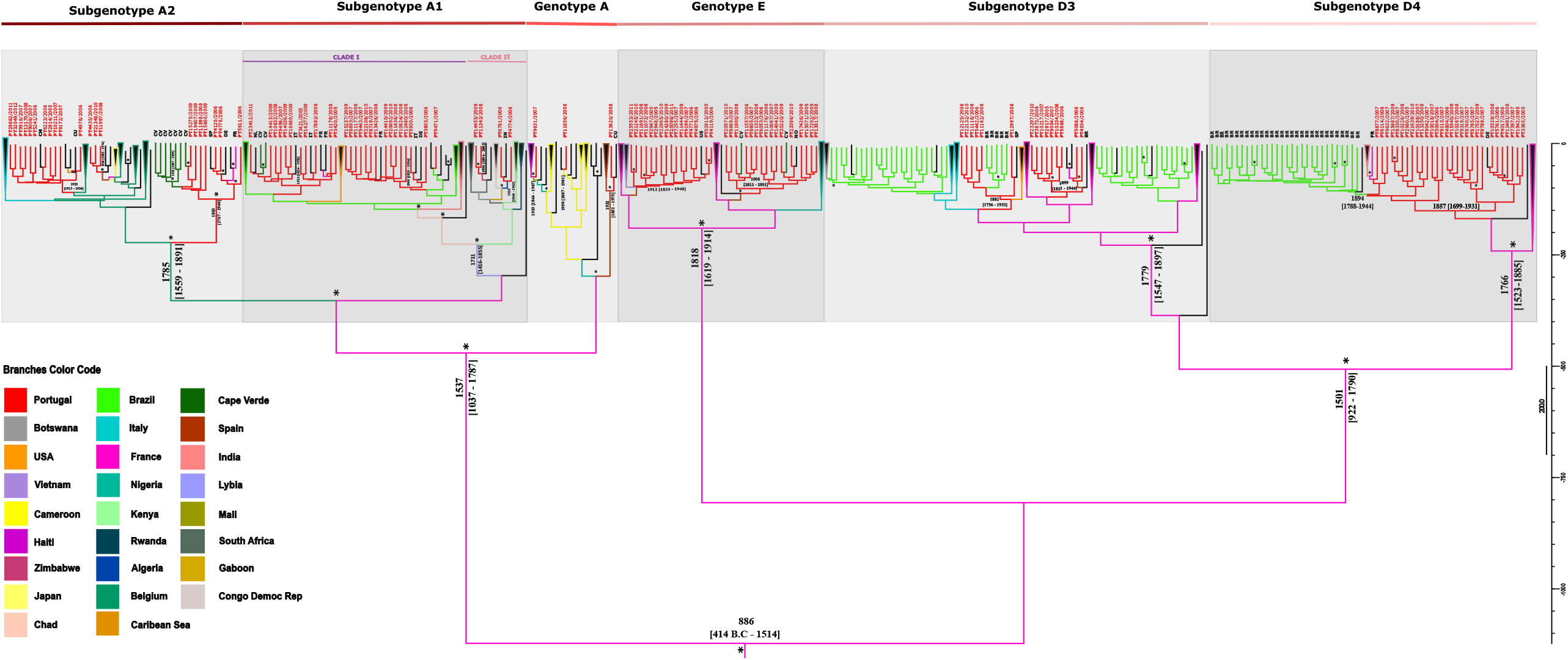
Bayesian maximum clade credibility tree of HBV partial polymerase gene sequences. The colored internal branches represent the locations of the parental nodes, as assigned by continuous Bayesian phylogeography (color code below the tree). Branches and tips of Portuguese isolates are labeled in red. External branches labeled with black initials represent the countries to where Portugal exported HBV strains (BR-Brazil; CM-Cameroon; CU-Cuba; CV-Cape Verde; DE-Germany; FR-France; IN-India; IT-Italy; NL-Netherlands; NO-Norway; SP-Spain; US-United States of America). The external branches representing Portuguese speaking countries, Brazil and Cape Verde, are highlighted in light green and dark green, respectively. Collapsed branches (triangles) represent clades that are not of primary interest for the study and are too large for displaying in full. They are filled in black because they include isolates from different origins, being outlined with the color of the country that originated them. Within subgenotype A1, Clade I – A1 Asian/American clade is labeled with a purple line; while Clade II – A1 African Clade is labeled with a rose line. The scale on the right of the tree represents years before the last sampling time (2012). Tree is annotated with genotypes/subgenotypes emergence dates and dates of introduction in Portugal. Branches with a posterior state probability support ≥0.9 are annotated with *. More information about control reference sequences (origin, date of collection and accession number) is available in figure S1 as well as in supplementary table 1.

Most of the HBV/A1 Portuguese strains cluster in A1 clade I (purple line) known as the Asian-American Clade, and only four Portuguese strains cluster together with the A1 clade II (rose line) known as the African Clade (Figure 1).

### Portugal exported HBV/A1 and HBV/A2 to several European countries and Cape Verde

Our results indicate that HBV/A diversified in North Africa around 1711 (95% HPD: 1416 - 1855) originating HBV/A1, and in Belgium near 1785 (95% HPD: 1559 - 1891) originating HBV/A2 (Figure 1; Table 3). Possibly, HBV/A1 had its origin in the region of Lybia and migrated across countries such as Chad. From this region, it seems to have been introduced in Brazil and India near 1869 (95% HPD: 1731– 1933). Finally, coming from Brazil, it was introduced in Portugal more than once around 1894 (95% HPD 1792 – 1942) and 1904 (95% HPD 1819 – 1945), later spreading from Portugal to other countries in Europe, mainly Italy and France, as well to Cape Verde Islands in Africa. A second route of A1 subgenotype introduction in Portugal occurred directly via Africa, possibly through Algeria and Zimbabwe around 1923 (95% HPD 1846 - 1963) and 1954 (95% HPD 1894 - 2002), respectively (Figure 2).

**Figure 2.**
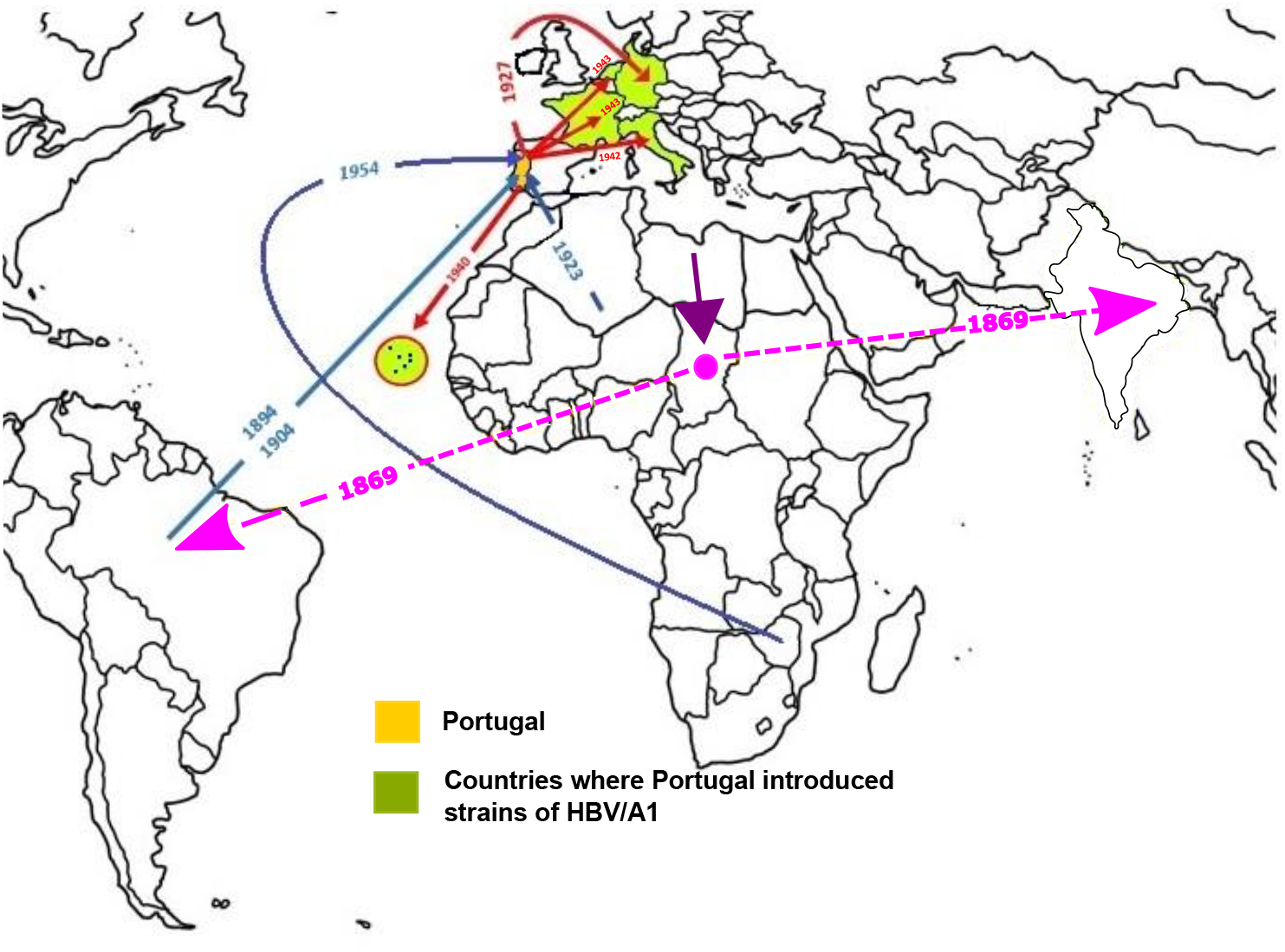
Probable major dispersal pathways of HBV/A1 as estimated by phylogeographic analysis. The blue arrows represent probable routes of introduction of the HBV/A1 in Portugal. The red arrows represent the probable dispersion paths of HBV/A1 from Portugal towards other countries. A significant dispersion event, prior to the introduction of A1 in Portugal, is represented in purple and pink: the purple arrow is located on the probable country of emergence of HBV/A1 indicating its dissemination direction; the pink circle represents the region from which HBV/A1 has been exported outside Africa. The dashed pink arrows simulate the likely route of exportation, that seemingly occurred simultaneously to Brazil and India.

In a different pathway, HBV/A2 has possibly emerged in Belgium between the middle of XVI and the end of XIX century, after migration from its original source in France (Figure 1; Table 3). After introduction from Belgium, HBV/A2 was established in Portugal in 1883 (95% HPD 1767 – 1940) and in 1925 (95% HPD 1913 - 1936) and spread not only to Spain, Germany, and France but also to Cuba and African countries such as Cameroon and Cape Verde. An additional HBV/A2 introduction in Portugal occurred around 1944 (95% HPD 1885 – 1974) via Spain (Figure 3).

**Figure 3.**
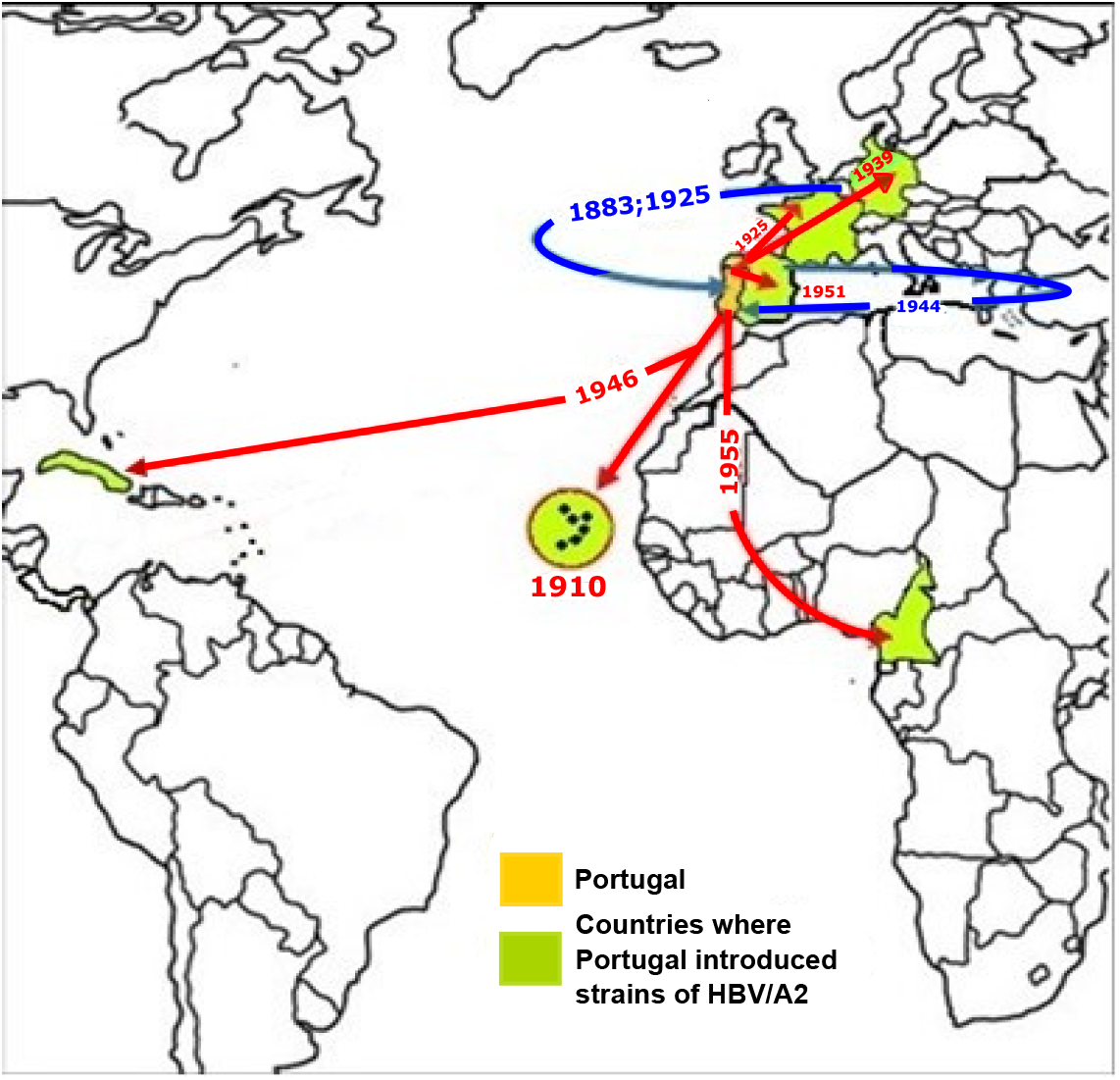
Probable major dispersal pathways of HBV/A2 as estimated by phylogeographic analysis. The blue arrows represent probable routes of introduction of the HBV/A2 in Portugal. The red arrows represent the probable dispersion paths of the HBV/A2 from Portugal towards other countries.

Our analysis revealed that the three Portuguese strains to which no subgenotype A classification was assigned have different origins. While one of them entered Portugal around 1932 (95% HPD: 1851 – 1975) via France and then Spain, the other two strains had an African origin. After divergence from the common ancestor, these strains seem to have diverged further in Nigeria, Cameroon, and Mali before finally being imported to Portugal. These last two strains were introduced in Portugal around 1933 (95% HPD: 1864 – 1967) and 1950 (95% HPD: 1887 – 1981) (Figure 1; Table 3).

### Portugal exported HBV/D4 to Brazil around 1894, causing a local epidemic

The common ancestor of D subgenotypes was dated to diverged in France around 1501 (95% HPD: 922 – 1790), branching off into subgenotypes D3 and D4 likely in the second half of XVIII century (Figure 1; Table 3). The introduction of HBV/D3 in Portugal occurred via France in two different occasions. It entered Portugal around the year of 1882 (95% HPD: 1756 - 1933) and then was exported to Brazil near 1909 (95% HPD: 1814 – 1952), while also spreading to USA and Spain (Figure 4_D3). The second importation to Portugal occurred near 1899 (95% HPD 1811 - 1944). These isolates were exported once again to Brazil and Spain. From this last country, they were later introduced again in Portugal from where, via France, they spread not only to Germany but also to Cuba (Figure 1). Other countries exporting HBV D3 to Brazil were France and Italy (Figure 4_D3; Figure 1).

**Figure 4.**
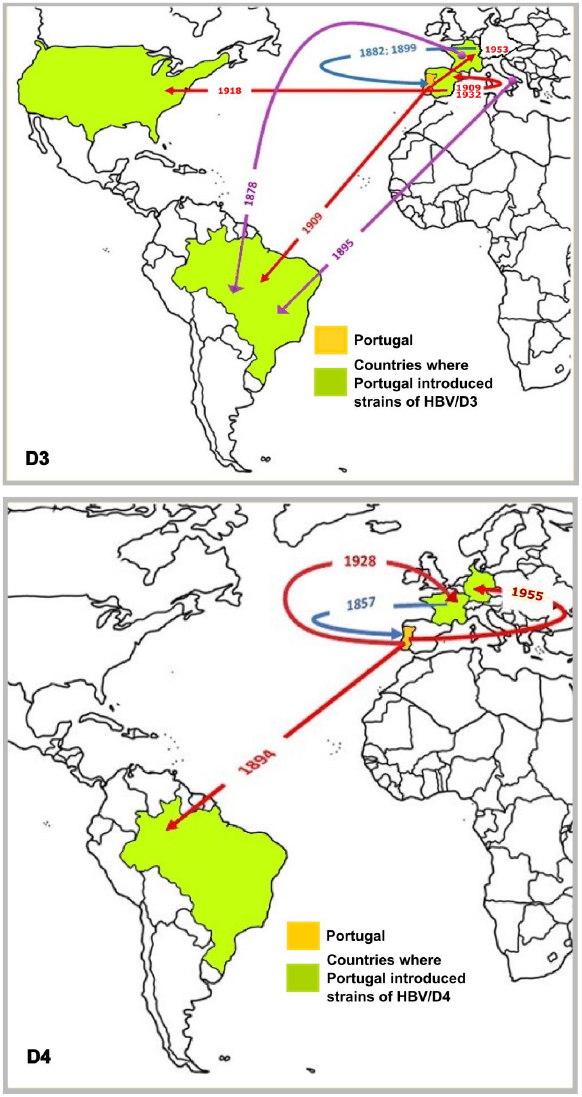
Probable major dispersal pathways of subgenotype HBV/D3 (above) and HBV/D4 (below) as estimated by phylogeographic analysis. The blue arrows represent probable routes of introduction of the subgenotypes in Portugal. The red arrows represent the probable dispersion paths of subgenotype from Portugal towards other countries. Purple arrows show the contribution of other countries, besides Portugal, to the introduction of D3 subgenotype in Brazil.

Subgenotype D4 originated in France, possibly entered Portugal near 1857 (95% HPD 1699 - 1931) from where it seems to have been exported to Brazil around 1894 (95% HPD 1788 – 1944), generating a cluster of Brazilian HBV/D4 strains. Portugal also exported HBV/D4 to France and Germany (Figure 4_D4; Figure 1).

### Genotype E was exported from Portugal to Cape Verde around 1940

Our data indicates that genotype E probably emerged more recently than all the others near 1818 (95% HPD: 1619 - 1914) in France (Figure 1; Table 3). After migrating to Spain, it was introduced in Portugal around 1906 (95% HPD: 1811 - 1951) and 1912 (95% HPD 1835 - 1948) creating two subclusters of Portuguese sequences (Figures 1 and 5). One of the clusters seems to be responsible for the exportation of HBV/E to Cape Verde and Norway region, while the other seems related to HBV/E spread to USA (Figures 1 and 5; Table 3).

**Figure 5.**
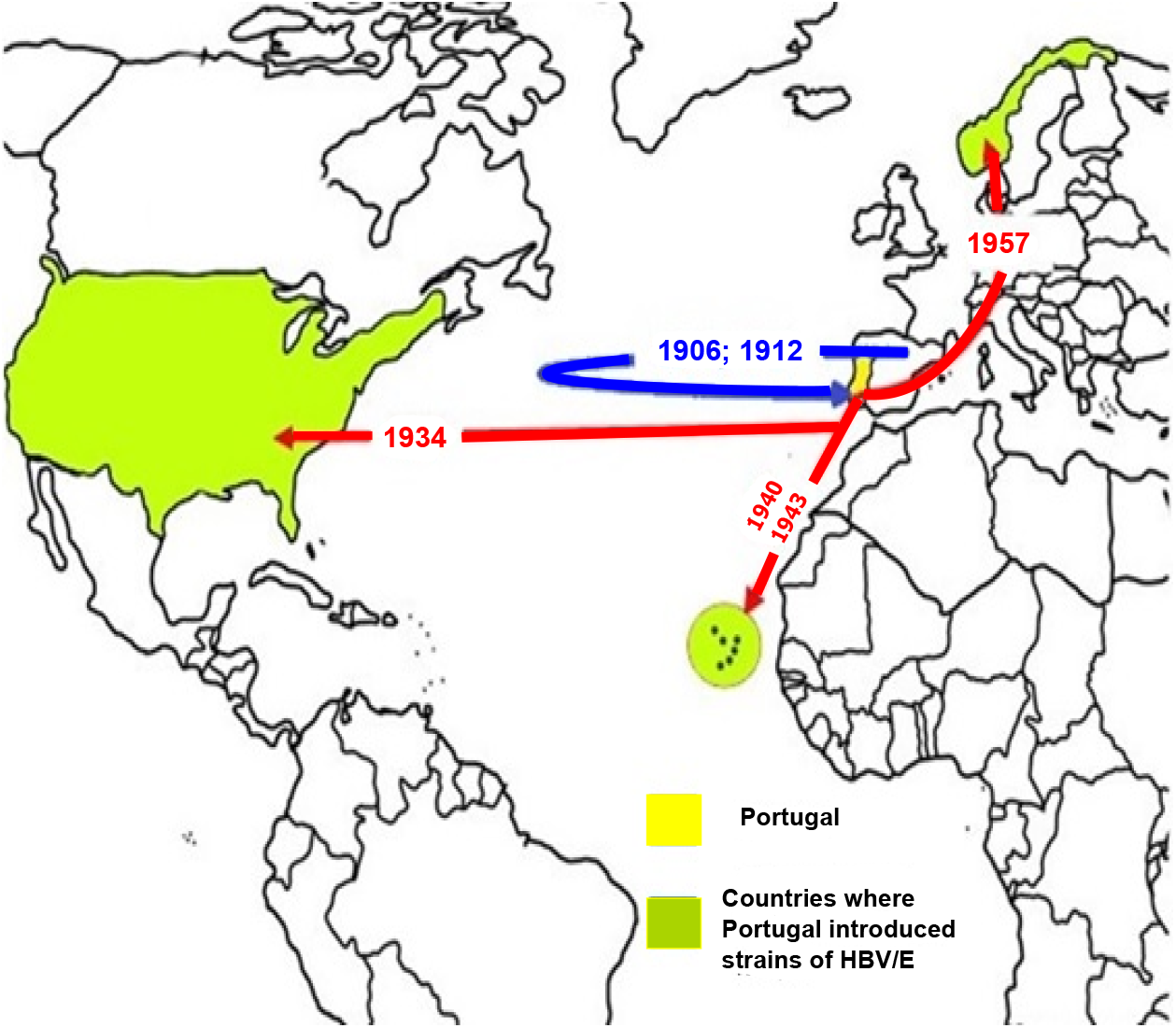
Probable major dispersal pathways of HBV/E as estimated by phylogeographic analysis. The blue arrows represent probable routes of introduction HBV/E in Portugal. The red arrows represent the probable dispersion paths of HBV/E from Portugal towards other countries.

## Discussion

To our knowledge, the present study is the first to reconstruct the spatial and temporal dynamics of HBV in Portugal using a Bayesian framework.

Earlier studies recurrently reported HBV genotype D as the most prevalent in Portugal, but most of them were done until 2008 or 2009 and those done most recently, until 2013 or 2014, were performed in the north or center of the country or with samples of a specific part of the population such as prisoners, pregnant women or blood donors (9, 15). Moreover, in none of these earlier papers was phylogenetic analysis used to classify the HBV strains. The current phylogenetic analysis, performed with HBV strains isolated from the general population followed in the major hospital of Lisbon between 2005 until 2012, revealed a higher percentage of genotype A strains compared to D, and genotype E as the least common genotype. This might reflect the changes in population migratory patterns in recent years, which particularly increased after the entry of Portugal in the European Union. During the period of samples collection in this study, there was a higher foreign influx of migrants from Cape Verde into Portugal, followed by Brazil and then Ukraine (22). HBV genotypes A (A1 and A2) and E prevail in Cape Verde (23). HBV/A1 is also the predominant subgenotype in Brazil, while HBV/A2 is the second most predominant subgenotype (24). This may explain the higher prevalence of genotype A than genotypes D in Portugal, when compared to earlier reports (15).

An estimated mean substitution rate of 9.45×10^−5^ s/s/y (95% HPD: 4.22×10^−5^ - 1.43×10^−4^) was obtained, which is in agreement with other recent HBV genome studies that also describe evolutionary rates between 10^−4^ - 10^−5^ s/s/y (24-27).

Molecular-clock-based analysis has been used to reconstruct the evolution of HBV as well as dating its origin, but the results have proven to be still unclear and controversial (28-30). Some studies have dated the origin of HBV as recent as 400 years ago (29-31) and other as old as more than 7000 years ago (32). Our data traced back the origin of HBV to the present region of France around the year 886 (medieval age) with a highest posterior density interval between the years 414 B.C (Ancient Age) and 1514 (Modern Age). In fact, there is recent evidence of the presence of HBV in Medieval and Neolithic human remains from Europe, proving that HBV is an ancient virus in this continent (32), and giving some support to the possibility that the origin of HBV may be in Europe. In agreement with Kramvis and Paraskevis (33), in our study the origin of HBV/A1 was traced back to Africa. However, contrarily to Kramvis’ suggestion that pointed to Southern Africa as origin of subgenotype A1, or even another theory that purposed East Africa as the origin (34), our data raises the hypothesis that the origin was actually in the North Africa. HBV/A1 evolution seems to have occurred not just along the North of the continent and Chad, from where it appears to have been exported directly to Brazil and India (clade I), but also in Kenya. From this region, it dispersed across Eastern, Central and Southern Africa (clade II), and then to Europe via Portugal and Belgium (see this detail in Figure S1_Supplementary material). These two distinct clades, correspond to the previously described Asian-American and African clades, respectively (33).

A previous study hypothesized that an alternative route to the slave trade could explain the fact that Brazilian HBV sequences clustered in the Asian-American clade instead of in the African clade, possibly being imported from East Africa or Asia by merchants in the middle of the XIX century (24). However, our results suggest instead, that HBV/A1 dispersed directly from Africa to both regions, Brazil, and India, in a period when Portugal was strongly present in both territories, probably contributing with African slaves as working labor to both continents. The African slave trade to Brazil, was especially from North Africa and Angola, to work in sugar cane plantations and gold mines. Slaves from Guinea were brought to Portugal where they were sold to other European kingdoms who also carried them to their colonies in Asia or South America (35). These facts raise the hypothesis that Portuguese slave trade was at the origin of the Asian-American clade. In addition, our temporal tree revealed that Brazil seemingly exported its strains to other American countries, namely Uruguay, Panama, Haiti, Cuba, USA and Argentina, while India exported its strains to Japan, China, Bangladesh and Philippines (see these details in Figure S1_Supplementary material). These historic liaisons provide a plausible explanation for the evolutionary proximity between HBV isolates from Americas and Asia.

Portugal seems to have imported most of the HBV/A1 strains from Brazil near the beginning of XX century, and then exported it to France, Italy, Netherlands, Germany and to Cape Verde, at this time still a Portuguese colony. In fact, the Portuguese role in exporting HBV/A1 into Cape Verde may explain why Cape Verdean HBV/A1 strains are more associated with the Asian-American clade, in particular with Brazilian strains, than with the African clade as should be expected (23).

HBV/A2 possibly emerged between the middle of XVI and the end of XIX centuries in Belgium, which seems to have played a main role in the spreading of HBV/A2 in Europe, exporting this subgenotype to Portugal more than once. A relevant role was also played by Portugal in spreading HBV/A2 outside of the European continent, mainly to Cape Verde between the beginning of the XIX and the middle of XX centuries. A2 strains from Cape Verde were previously described to group into a single cluster quite separate from the HBV/A2 strains of the other geographic regions (continental Africa, Asia, the Americas or Europe), with the exception of a strain from Poland (23), a European country to which Portugal also seemingly exported HBV/A2 via France.

Our results provide the first indication that Portugal was the source of HBV/A2 isolates found in Cape Verde.

Providing further support to this, we found two common genomic regions of Cape Verdean and Portuguese sequences that were different from those observed in sequences from other regions of the world. Amino acid V588, present in all the sequences of both countries that clustered together, appeared just in 7% of the remaining foreign sequences. H606 was present in all the sequences from other regions, while Cape Verde and Portuguese sequences presented mainly D and N at this position (data not shown).

Evidence from our Bayesian analysis suggests that HBV/D emerged earlier than HBV/A, around the year of 1501, with a confidence interval that covers the vast period from 922 to 1790. Our study points to a putative origin of HBV/D in France, while previous phylogeographic analysis indicated North Africa/Middle East (36), Southern Europe, Central/Eastern Europe, Syria or Martinique (25) as the putative origin of HBV/D. The lack of HBV/D sequences from more diversified locations in the world and from consistent genomic regions is probably limiting consensus conclusions in phylogeographic studies.

Our results indicate that Portugal played a fundamental role in the dissemination of HBV/D3 and HBV/D4 during the period that covers a great wave of Portuguese emigration to the American continent in the late XIX and early XX centuries. During this period, there were great human migrations in different parts of the globe, but none can compare with the 44 to 52 million Europeans that crossed the Atlantic to the American continent between 1815 and 1914 (37). This was caused by the European economic crisis and the labor crisis caused by the progressive cessation of slave traffic worldwide during the second half of XIX century. Many Portuguese from the continental region emigrated to Brazil, not only because of the language but also for religious affinity. On the other hand, Portuguese from Azores emigrated mostly to the United States (37).

All Portuguese HBV/D4 sequences clustered together suggesting a single introduction of this subgenotype in our country via France. This Portuguese cluster seems to be the most likely and almost exclusive origin of HBV/D4 infections in Brazil, whose sequences also cluster monophyletically with just one exception. This clustering effect of D4 Brazilian sequences was previously described, and a single introduction of D4 in Brazil had been proposed by the authors with a probable time of introduction of 1848 (CI: 1062-1946) (25). However, spatial origin was pointed at Martinique, which the authors considered incompatible with epidemiological and historical data of HBV in the Americas (25). Our results help to further clarify these previous results, now showing that Portugal is at the origin of Brazilian D4 strains with its most recent common ancestor (tMRCA) dating around 1894 (CI:1788 to 1944) which coincides with the period of massive emigration from Portugal to Brazil by the end of XIX century (37). As the confidence interval still covers the end of the XVIII century, it is not possible to exclude the slave trade factor contributing for the introduction of this subgenotype in Brazil. However, the parallel exportation of HBV/D4 from Portugal to France (1928) and Germany (1955), which were secondary emigration destination of Portuguese people in the first half of last century, further support a later exportation to Brazil and therefore the theory of Portuguese emigration.

Subgenotype D3, introduced in Portugal perhaps a few years later than the HBV/D4 (late of XIX century) but also with a probable origin in France, seems to have been exported with Portuguese population together with the Italians and French to Brazil in the beginning of the XX century, around 1909 (1814 to 1952). HBV/D3 was previously described to have been introduced in Brazil around 1799 (1615 to 1976) by Southern European people (25), especially during the mass European emigration from different countries encouraged by the Brazilian government campaigns, such as Italians, Portuguese, Spanish, Japanese, Germans, Lebanese and Syrians (38). The Italians, who were divided between emigration to Europe and Argentina, only came to Brazil in large groups in the last two decades of the XIX century (37), probably introducing genotype D3 in the country as previously proposed (24). Interestingly, this agrees with our results that indicate that the tMRCA is near 1895 for a group of HBV/D3 Brazilian sequences with origin in an Italian cluster. From the moment the Portuguese people first arrived in South American lands there was always a flow of Portuguese people to the region that became Brazil. Nonetheless, the largest flow of Portuguese emigration started in late XIX and the beginning of XX centuries (37), which agrees with our estimated tMRCA of 1909 for the introduction of Portuguese HBV/D3 in Brazil. The confidence interval of this tMRCA has a lower limit of 1814, which is already very close to Brazil’s independence (in 1822) and consequently to the end of the slave trade by the Portuguese people. This gives more support to the possibility of HBV/D3 introduction in Brazil by the Portuguese to have occurred during the emigration flow in XIX - XX centuries rather than with the slave trade.

Although with less intensity, Portugal also seems to have introduced HBV/D3 subgenotype in the USA, Spain and France, which agrees with other secondary migratory destinations of Portuguese people (37).

Genotype E has been assumed to have a more recent origin probably in the present region of Nigeria around the last 130 to 200 years, which is supported by its low genetic variability characteristic of a shorter evolutionary history (39, 40). The tMRCA found in our study, 1818, is in full agreement with this. However, contrarily to the theories of those authors, our data supports France as the origin location for HBV/E instead of Africa. One possibility for this disparity is the fact that some HBV/E sequences available in the databases don’t indicate sample collection date and geographical origin and therefore could not be used in our study.

Despite the low genetic variability, HBV/E is hyperendemic throughout West Africa, reason why it has been proposed a rapid spread in countries along the West African coast from Guinea to the Central African Republic and subsequently towards Eastern and Southern countries, such as Sudan, Angola and the Democratic Republic of the Congo (39, 40). As revised by Muller *et al* in 2009, an explanation for this phenomenon would be iatrogenic transmission in mass-injection campaigns with unsafe injections by the colonial governments (41). Interestingly, Portugal seems to have imported HBV/E from Spain in the beginning of XX century. Our estimated dates of introduction of HBV/E in Portugal fall precisely within the period between the 1880s and 1930s, previously described as the most significant expansion period of this genotype (40). While the Portuguese role was markedly important for the spread of the HBV/A1 and HBV/A2 during the slave trade, a clear difference is evident in this case since Portuguese sequences are not causing outstanding exportations to other countries. This is in agreement with the absence of HBV/E in descendants of African slaves in South America that was assumed to indicate that this genotype could not have been present in the West African population in the period of slave trade (40, 42, 43). The absence of the HBV/E from the South American continent also indicates that Europeans who emigrated to the Americas did not introduce it there. From our point of view, this fact reinforces the idea that during the period of emigration from Europe to the American continent, in the end of the XIX century and the beginning of the XX century, HBV/E was probably still absent or weakly incident in Europe. This explanation is also in line with the reduced Portuguese exportation of HBV/E to other countries.

The main limitation of this study was the inevitable exclusion of the analysis of sequences that were present in databases but that did not gather all the necessary conditions to be used in the spatiotemporal reconstruction. Many sequences corresponding to the region of the *pol* gene under analysis did not have a date of sample collection or country of origin and had to be excluded. For this reason, some regions of the globe were underrepresented, limiting the analysis. Some regions such as Angola, Guinea Bissau, Mozambique with which Portugal maintained a close relationship were not equally represented in the datasets of all the studied genotypes.

## Conclusions

HBV genotype A, D and E prevail in Portugal. HBV/D4 was the first subgenotype to be introduced in Portugal around 1857 (HPD 95% 1699 - 1931) followed by subgenotypes D3 and A2 a few decades later, and genotypes E and A1 more recently. Portugal had a major role in the dispersion of HBV to Africa, America, and Asia. This work provides new insights in the early and global epidemic history of HBV.

## Data Availability

All data produced in the present study are available upon reasonable request to the authors

## Figures subtitles

**Figure S1. Bayesian maximum clade credibility tree of HBV partial polymerase gene sequences**. The colored internal branches represent the locations of the parental nodes, as assigned by continuous Bayesian phylogeography (color code on the left bottom of the tree). External branches are labeled with the accession number, isolates origin country and year of sample collection (more information can be accessed in supplementary table 1). Countries to where Portugal exported HBV strains are labeled in blue. Branches and tips of Portuguese isolates are colored/labeled in red. The external branches representing Portuguese speaking countries, Brazil and Cape Verde, are colored in light green and dark green, respectively. The scale at the bottom of the tree represents years before the last sampling time (2012). Within subgenotype A1, Clade I (A1 Asian/American clade) is labeled with a purple line, while Clade II (A1 African clade) is labeled with a rose line. Tree is annotated with the emergence dates of genotypes/subgenotypes and dates of their introduction in Portugal. Branches with a posterior state probability support ≥0.9 are annotated with *.

**Table S1**. HBV Control Sequence details used for the spatio-temporal analysis: origin country, collection date and accession number. The number of sequences by genotype, country and date of collection can be checked by filtering.

## Acknowledgements

This work was performed in the context of a PhD study, whose student’s fellowship (SFRH/BD/99507/2014) was supported by Fundação para a Ciência e Tecnologia (FCT), POCH program, Portugal 2020 and European Union/Social European Fund (FSE).

This work was supported by FCT through funds to GHTM-UID/Multi/04413/2013 and GHTM-UID/04413/2020.

This work was supported by the Portuguese Agency for Scientific Research, “Fundação para a Ciência e a Tecnologia” (FCT) through projects UIDB/04138/2020 and UIDP/04138/2020.

