## Supplementary figures and images for "Phylogeography of hepatitis B: The role of Portugal in the early dissemination of HBV worldwide"

### Figure S1

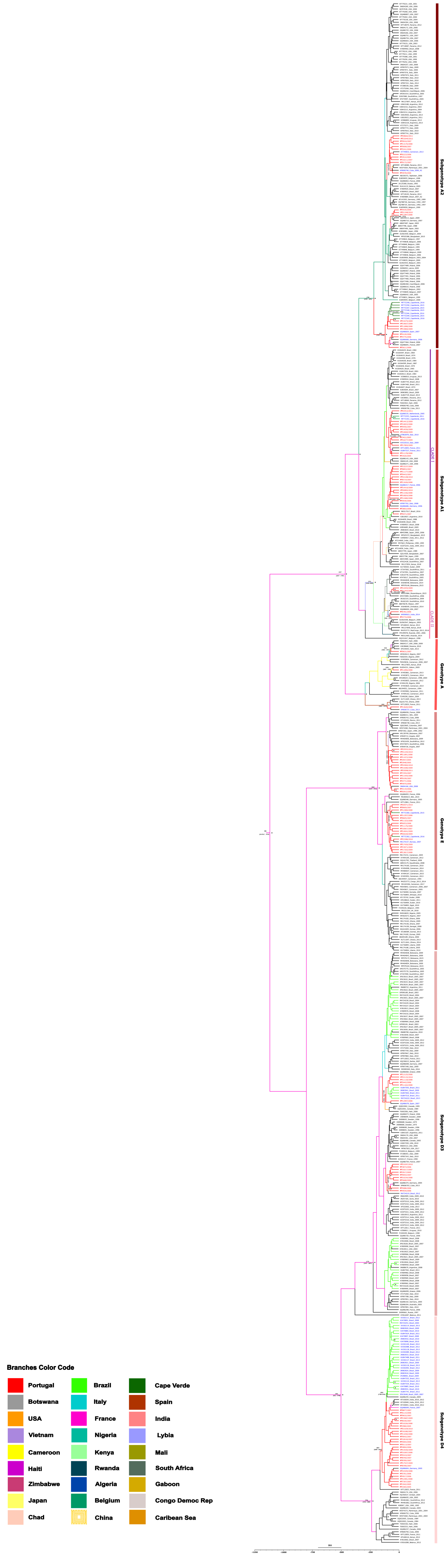
